# Reconstructing unseen transmission events to infer dengue dynamics from viral sequences

**DOI:** 10.1101/2020.11.25.20236927

**Authors:** Henrik Salje, Amy Wesolowski, Tyler Brown, Mathew Kiang, Irina Maljkovic Berry, Noemie Lefrancq, Stefan Fernandez, Richard Jarman, Kriangsak Ruchusatsawat, Sopon Iamsirithaworn, Warunee P. Vandepitte, Piyarat Suntarattiwong, Chonticha Klungthong, Butsaya Thaisomboonsuk, Kenth Engo-Monsen, Caroline Buckee, Simon Cauchemez, Derek Cummings

**Affiliations:** Department of Genetics, University of Cambridge, Cambridge, UK; Mathematical Modelling of Infectious Diseases Unit, Institut Pasteur, UMR2000, CNRS, Paris, France; Department of Biology, University of Florida, Gainesville, Florida, USA; Harvard University, Cambridge, USA; Center for Population Health Sciences, Stanford University School of Medicine, Stanford, California USA; Viral Diseases Branch, Walter Reed Army Institute of Research, Silver Spring, USA; Department of Virology, Armed Forces Research Institute of Medical Sciences, Bangkok, Thailand; National Institute of Health, Department of Medical Sciences, Ministry of Public Health, Nonthaburi, Thailand; Department of Disease Control, Ministry of Public Health, Nonthaburi, Thailand; Queen Sirikit National Institute of Child Health, Bangkok, Thailand; Telenor, Oslo, Norway; Emerging Pathogens Institute, University of Florida, USA

## Abstract

For most pathogens, transmission is driven by interactions between the behaviours of infectious individuals, the behaviours of the wider population, the local environment, and immunity. Phylogeographic approaches are currently unable to disentangle the relative effects of these competing factors. We develop a spatiotemporally structured phylogenetic framework that reconstructs transmission across spatial scales and apply it to geocoded dengue virus sequences from Thailand (N=726 over 18 years). We find infected individuals spend 96% of their daytime hours in their home community compared to 76% for the susceptible population (mainly children) and 42% for adults. Dynamic pockets of local immunity make transmission more likely in places with high heterotypic immunity and less likely where high homotypic immunity exists. Age-dependent mixing of individuals and vector distributions are not important in determining transmission. This approach provides previously unknown insights into one of the most complex disease systems known and will be applicable across pathogens.

---

As with other endemic pathogens, widespread, sustained co-circulation of dengue viruses (DENV), effectively masks the dynamics of individual lineages^1–3^. The co-occurrence of unrelated transmission chains means we still only have a limited understanding of how DENV spreads, including the role for human mobility of both infected individuals and the surrounding susceptible population, age-specific mixing, and local heterogeneities in serotype-specific population immunity and mosquito density. These mechanistic knowledge gaps help explain our failures to control a pathogen that continues to cause 50 million annual symptomatic infections globally^4^. The use of pathogen sequences has the potential to help. However, existing phylogeographical approaches can only provide limited mechanistic insight into drivers of spread as they focus on rates of flow between locations present in a phylogeny, based on assumptions of mass action and using traits attached to the observed sequences (i.e., cases) only, which in the case of DENV will only ever represent <1% of all infections^5–7^. Critically, these approaches do not consider that viral flow is made up of sequential transmission events with each event arising from a complex interplay of individual-, population-, environmental-, and viral-level factors. Further, the bulk of available sequences typically come from a few locations with most locations providing no data. Current phylogeographic approaches will infer a viral flow between observed locations without consideration that transmission events that link two observed sequences will be unobserved and often in unsampled locations.

Here we develop an inference framework that fills this knowledge gap by explicitly considering individual transmission events. By using the generation time distribution for dengue (Figure S1), we derive estimates of the number of generations that separate each pair of sequences in a time-resolved phylogeny and consider viral mobility for a single transmission generation. This shift of focus to single transmission generations, rather than overall viral flow, allows us to develop detailed mechanistic models of how viruses are moving at a tractable and interpretable scale. For example, we separately model population movement for infected individuals, the susceptible population (mainly children) as compared to adults, allowing for transmission to occur in the infector’s community, in the infectee’s community or in a tertiary location. We assume the local scale of movement of the *Aedes* mosquito means only human mobility can drive spread between locations^8^. We allow for the disabling symptoms from dengue to result in reduced mobility in cases compared to the susceptible population^9^; that transmission may occur in an age-structured manner^3^; and that the spatial heterogeneity in vector distributions and the dynamic nature of local serotype-specific population immunity may affect where successful transmissions occur^10^. Using the transmission probabilities for a single generation, we probabilistically integrate over all possible pathways for the total number of transmission generations that link the observed locations for each pair of sequences, thereby capturing movement in unsampled locations. In parallel, we incorporate the probability of sequencing (i.e., observing) an infection at each space-time unit, thereby explicitly incorporating space-time biases in sampling. We fit our models in a maximum likelihood framework that incorporates uncertainty from the evolutionary processes, generation time distribution, and sampling using a bootstrap approach.

We apply our framework to dengue in Thailand using 726 sequences obtained from seven different provinces sampled over an 18 year period (1995 to 2012) (Figure 1A,D), from which we build time-resolved phylogenies (Figures 1E-H)^11^. In Bangkok, the home location of the cases were also geocoded (N=467) (Figure 1B). To inform our model, we use data from the third largest mobile phone operator in Thailand that empirically captures how adults move (N=11.4 million subscribers; 26% market share), estimated distributions of vector presence (Figure S2) and the long-term spatio-temporal distribution of serotypes (Figure 1C)^12^. We explore viral movement over two different spatial scales: within central Bangkok (N=337 1km^2^ grid cells throughout the center of the city) and nationwide (N=76 provinces with a mean area of 6,700km^2^ each).

**Figure 1.**
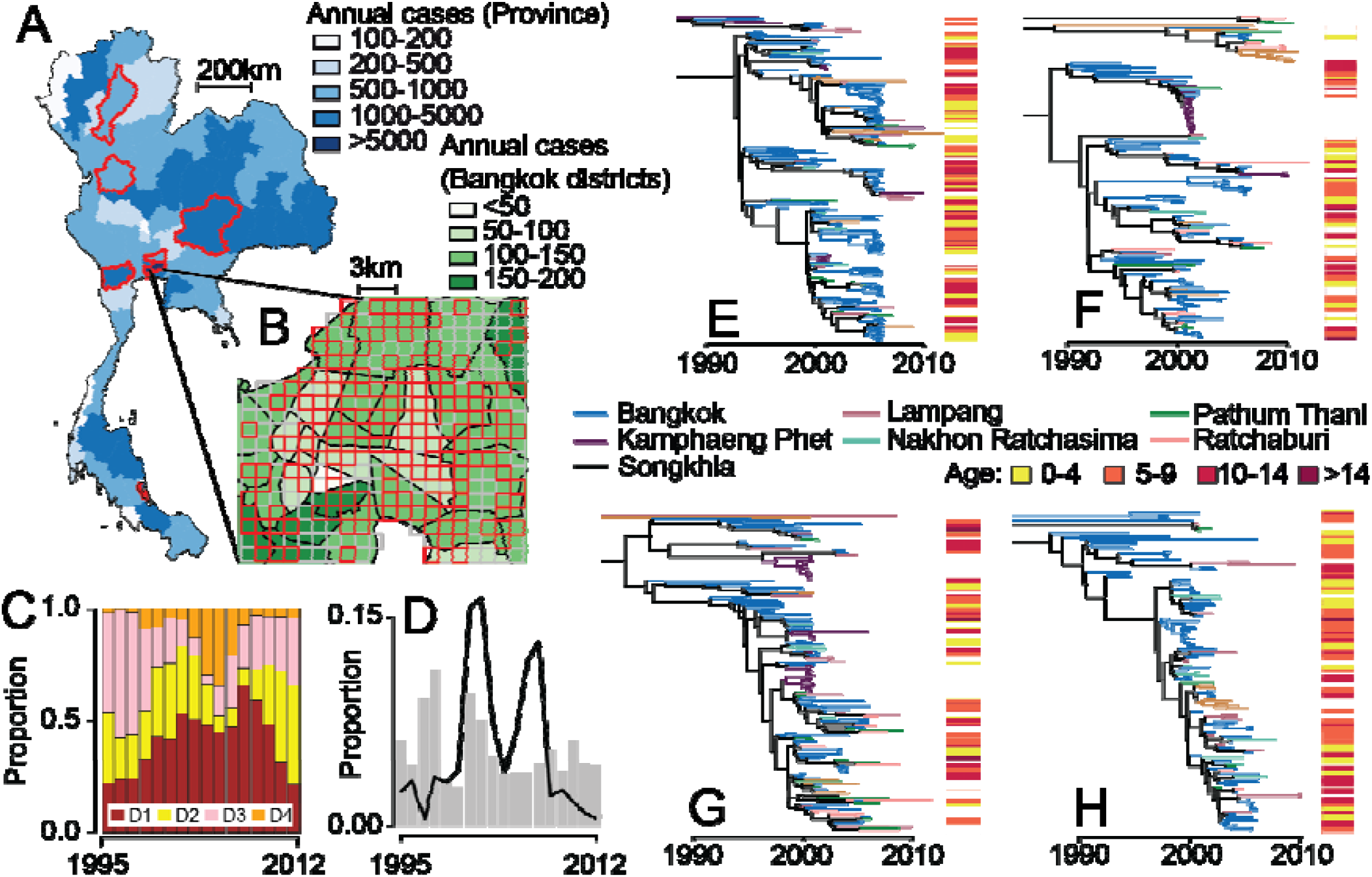
**(A)** Mean annual reported cases per province. The provinces in red represent sentinel sites where sequenced viruses are available. **(B)** Mean annual reported cases per district in central Bangkok. The grid cells in red are locations with available sequences. **(C)** Serotype distribution by year. **(D)** The distribution of all reported cases (grey) and available sequences (black line) from 1995-2012 by the year they were reported. Time resolved phylogenetic trees for **(E)** DENV1, **(F)** DENV2, **(G)** DENV3 and **(H)** DENV4 with location and age information.

Using our framework, we estimate that in Bangkok, infected individuals spend 96% (95% CI: 88%-100%) of daytime hours within their home cell, compared to 76% (95% CI; 42%-86%) for susceptible individuals and 42% for adults (95% CI; 41%-43%) (Figure 2A, Figures S3-4). These differences likely reflect that susceptibility is concentrated in children, who may spend more time near their home than adults. It also suggests that while subclinical DENV infections are common, they typically still result in severe enough symptoms to change daily routine and limit mobility. Importantly, these estimates of differential mobility hold for the intervening unseen transmission events, as well as the observed cases in the phylogeny. These patterns of differential mobility are also observed at the national scale, with cases spending 96% of the time within their home province (95% CI: 95%-100%), compared to 95% for the susceptible population (95% CI: 92%-100%) and 87% for adults (95% CI: 86%-88%) (Figure 2B). Local serotype-specific immunity also appears important, with transmission more likely to occur in places that have seen increases of other (heterotypic) serotypes circulating in the previous two years and less likely to occur in places with increased cases of the same (homotypic) serotype within the same timeframe (Figure 2D-E, Figures S3-4). However, the overall incidence of reported cases in the home cell or province of the susceptible population is not associated with differences in transmission risk, highlighting the complex relationship between observed case incidence and underlying infection risk. Further, the probability of *Aedes aegypti* presence is not linked to transmission risk (Figure 2F, Figures S3-4), although, for Bangkok, this may be driven by limited heterogeneity in estimated presence across the city (Figure S2). We also find no evidence of age-dependent transmission, with 0.12 (95%CI: 0.09-0.42) of transmissions in children occurring between individuals of <2y in age-difference, which is not statistically different to models with no age-structure in transmission and significantly less than estimates derived from empirical measurements of age-mixing in populations^13^, suggesting that the intermediary vector removes the effect of assortative mixing (Figure 2G, Figure S5).

**Figure 2.**
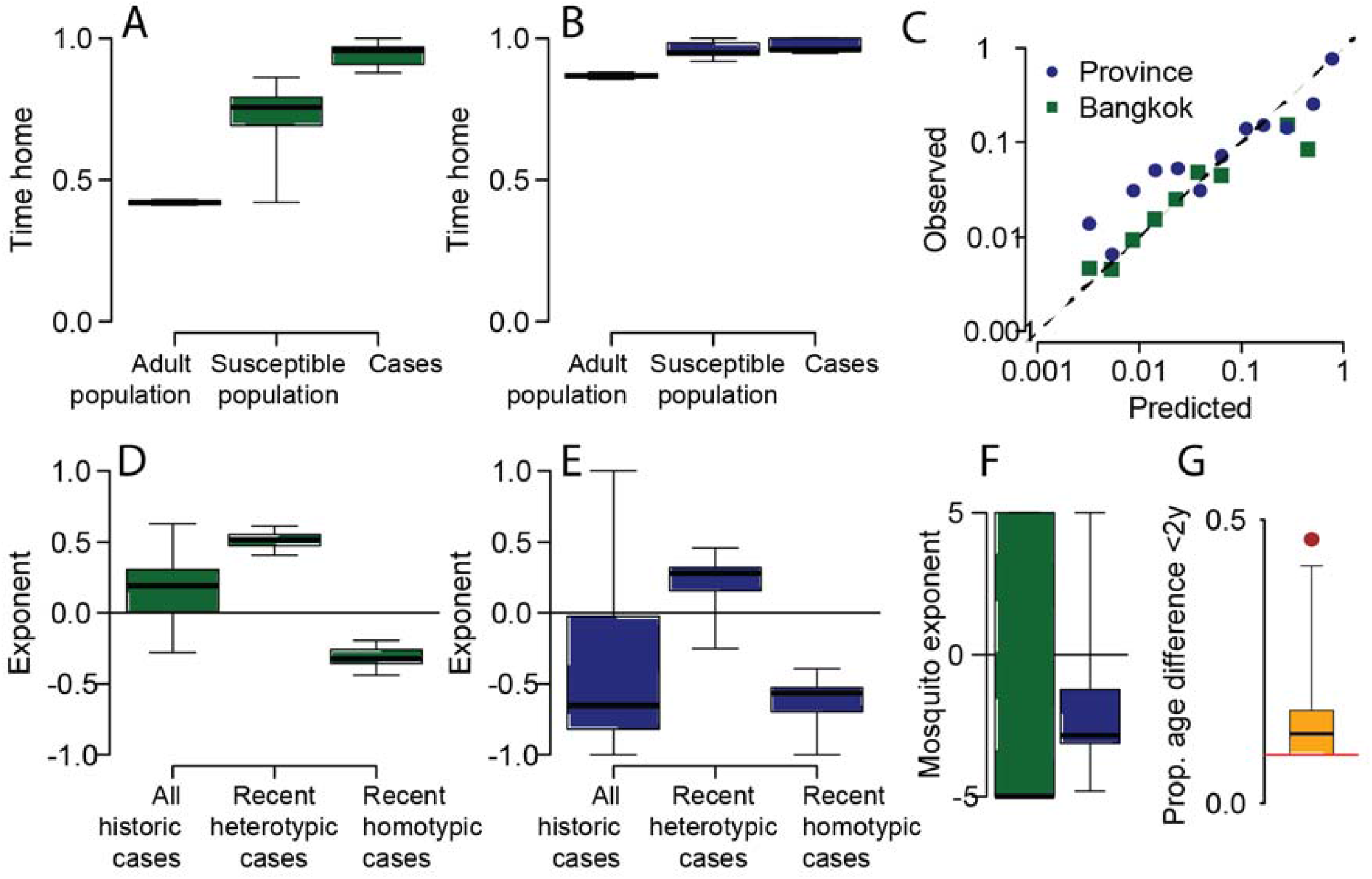
**(A)** Estimated proportion of daylight hours spent within your home 1km2 grid cell in Bangkok for adults, susceptible individuals and cases. **(B)** Estimated time within your home province. **(C)** Comparison of predicted and actual observed location of viruses from 20% held out locations (where viruses from held out locations are not included in the model fitting process). **(D)-(E)** Impact of overall number of cases throughout study period and recent (defined as in the previous 1-2 years) homotypic and heterotypic cases on the flow of virus to any location at any time. **(F)** Impact of estimated probability of *Aedes aegypti* occurence on probability of infection within any location (green - Bangkok, blue - provinces). **(G)** Proportion of infector-infectee pairs being within 2y in age. The red line represents homogeneous mixing. The red circle represents the estimate using age-mixing patterns from POLYMOD.

Our model is able to accurately estimate the probability of observing viruses in held out locations, both within Bangkok and at the nationwide scale (correlation between observed and estimated locations of held out sequences of 0.94 in Bangkok and 0.94 nationwide) (Figure 2C). This strongly suggests our framework can characterize viral movement in unobserved locations and highlights how non-representative sequencing approaches can still provide accurate descriptions of overall virus mobility. However, sampling biases do need to be explicitly incorporated as assuming unbiased observation results in very different parameter estimates, including falsely high estimates of between-location population movement (Figure S6). Using a simulation framework, we show we are able to accurately recover known parameter values, even under biased observation (Figures S7).

We use our fitted model to characterize the movement of the virus at each transmission generation. We find that if a virus is introduced into a randomly selected province, it is on average 4.3 times as likely to have travelled to Bangkok after a single transmission generation compared to anywhere else (95%CI: 2.4-7.0) and 11.4 times as likely to be in the capital after 20 generations (equivalent to approximately one year of sequential transmissions) (95%CI: 6.3-19.4) (Figures 3A, S8). The flow to larger cities is not restricted to Bangkok. with the likeliest destinations after 20 generations also being where the largest population centers are located (Figure 3A). Substantial heterogeneity is also observed at the local scale, with the virus tending to go to the hyper-urban city center in Bangkok (Figures 3B). Overall, within Bangkok, we find that 34% of infections occur outside the 1km^2^ home grid cell of an infected individual (95%CI: 26%-43%). This is compared to 4% of individual case mobility being outside cells (Figure 3C). After 20 generations, only 2.6% of viruses are still within the same Bangkok cell and 34% are within the same province (Figure 3D).

**Figure 3.**
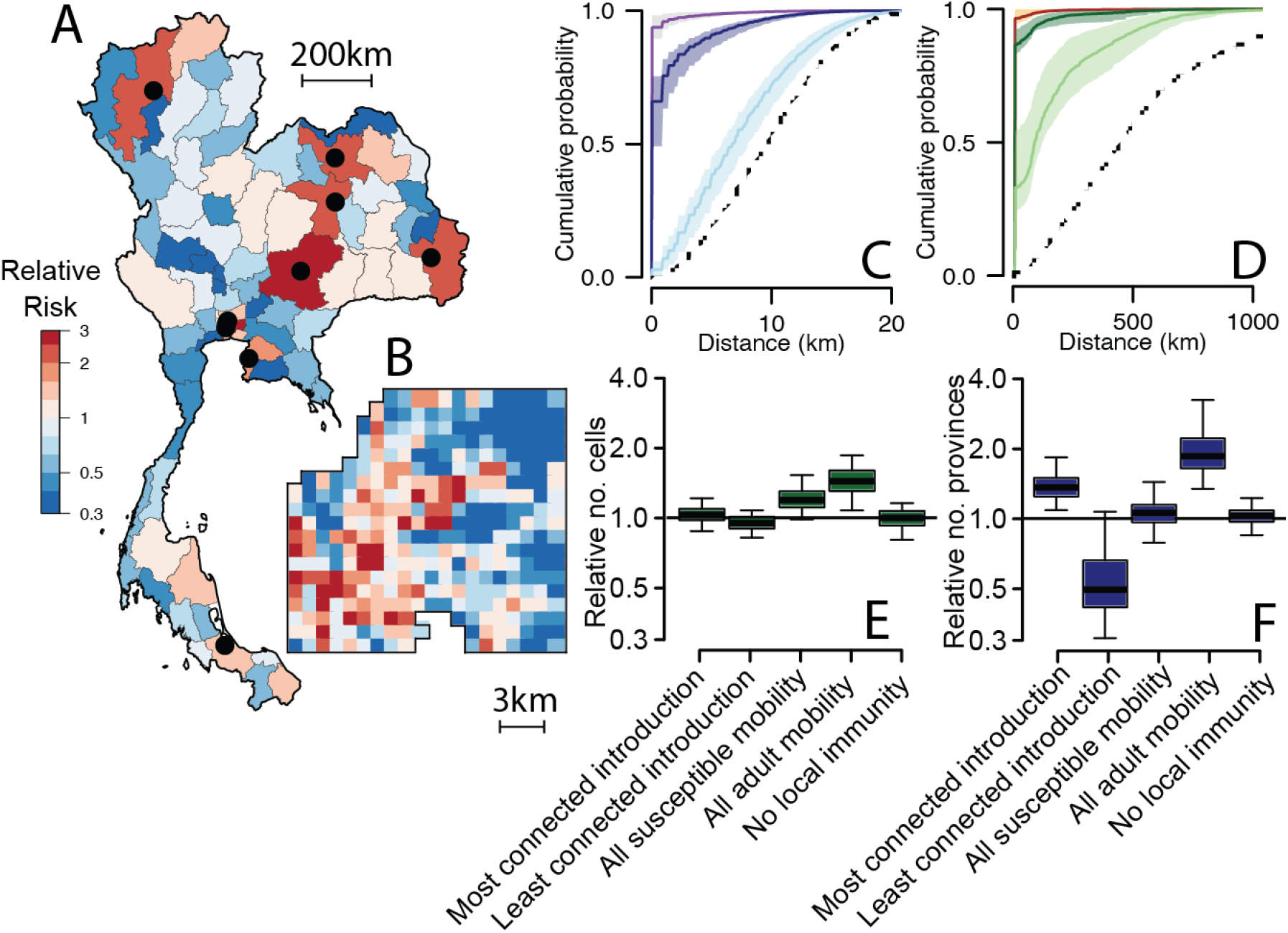
**(A)** Relative risk of movement of virus to each province compared to moving to a randomly selected province after 20 generations. The black dots represent the 10 largest cities in Thailand. **(B)** Relative risk of movement of virus to each grid cell within central Bangkok after 20 generations compared to moving to a randomly selected cell. The cumulative distance distribution for national **(C)** and central Bangkok **(D)** of how far cases are from home (purple/brown), the distances of single transmissions (dark blue/dark green) and distances after 20 transmissions (light blue/light green). The dashed line represents the cumulative distribution function of completely spatially random movement. **(E)** The number of Bangkok locations with at least one case for different scenarios relative to that in the base model after 20 generations. The different scenarios are initial introductions are in the most connected location, initial introductions are in the least connected location; no reduced mobility in cases compared to the susceptible population; case/susceptible population mobility is equal to adult mobility; no impact of local immunity on where transmission occurs. **(F)** For the same scenarios, the number of provinces with at least one case relative to that in the base model after 20 generations.

While most transmissions occur within the home cell of an infected individual, we find that when transmissions do occur further away, local heterogeneity in patterns of serotype-specific population immunity means that the pathway taken by viruses depends on the serotype (Figure S9). Within Bangkok, on average, 85% (95%CI: 81-89%) of the likeliest location after a single transmission event was the same across serotypes dropping to only 44% (95%CI: 38-53%) overlap after 20 generations. These effects are not observed at a larger scale, where the likeliest destination province remained largely the same across serotypes.

Using our estimates of viral movement, we build a stochastic simulation study to estimate the time it takes for viruses to spread across Bangkok and across the country assuming introductions in randomly selected locations. We find that after 20 transmission generations the virus will be in 27% of all provinces (95%CI 15-38) and in 32% of cells within central Bangkok (95%CI: 25-43) (Figure S10). We find that local immunity and the reduced mobility of cases compared to the susceptible population has minimal effect on the number of locations affected, however, if the mobility of susceptible individuals matched that of the adult population, there would be 1.9 times as many infected provinces (95%CI: 1.4-3.3), with a similar effect at the local Bangkok scale (RR: 1.5, 95%CI: 1.1-1.9) (Figure 3E-F). For arboviruses, such as Zika and chikungunya viruses, where limited immunity means most infections are in adults, we could therefore assume a more rapid dispersal of the virus compared to DENV^14^. We observe consistent patterns across different effective reproductive numbers (Figure S10).

By explicitly characterising the mechanisms of individual transmission generations and integrating the mobility of populations, our framework brings inference to a tractable scale, and allows unbiased inferences to be made despite minimal and heavily biased sequence availability. Individual transmission generations are also those most relevant for targeted interventions and can help predict future flows. While we have used this framework for DENV, it is applicable to any pathogen phylogeny where the generation time distribution is known and there exists spatial information or other discrete traits.

## Data Availability

All sequences are available from GenBank. The accession numbers are set out in Table S1.

## Disclaimer

Material has been reviewed by the Walter Reed Army Institute of Research. There is no objection to its presentation and/or publication. The opinions or assertions contained herein are the private views of the author, and are not to be construed as official, or as reflecting true views of the Department of the Army or the Department of Defense.

## Author contributions

HS developed the method, conducted the analysis and wrote the first draft of the paper. SC, AW and DATC helped methods development. TB, MK, IMB, SF, RJ, KR, SI, WV, PS, CK, BT, KE-M, CB worked on obtaining data for the analyses. All authors contributed to revising the paper.

## Methods

### Data sources

#### Sequence data and associated metadata

We use all available full genome sequence data from all four serotypes from Thailand covering the period 1994 and 2012 where province level spatial information is available. All sequences are available from GenBank. The accession numbers are set out in Table S1. For sequences from Bangkok, we also have household coordinates for a subset of 432 sequences. In Bangkok, the sequences come from Queen Sirikit National Institute of Child Health, a large children’s tertiary care hospital based in the center of the city. Outside Bangkok, most sequences come from the national surveillance system of dengue run by the Ministry of Public Health. They perform confirmatory testing and viral isolation and sequencing of samples from sentinel hospitals based around the country. The hospitals are based in the following five provinces: Lampang, Ratchaburi, Songkhla, Pathum Thani, Nakhon Ratchasima. In addition, there are sequences available on GenBank from Kamphaeng Phet province.

#### Case data

All cases of dengue are notifiable and are reported to the Thai Ministry of Public Health. We extract the number of cases per year for each year and each of the 76 provinces in the country between 1994 and 2012. To estimate serotype-specific case data in each year we used the serotype distributions of geocoded cases from QSNICH for Bangkok (N=11,583 cases). For the rest of the country, we used serotype-specific case data from the Ministry of Public Health. Outside the five sentinel surveillance sites, this data mainly comes from ad hoc samples sent to the ministry for testing. Altogether this represents a serotype-specific database of 27,586 cases covering 67 of the 76 provinces. For each province and year, we calculated the proportion of cases that were caused by each serotype. Where there were no samples from that province-year, we used data from the closest province in that same time period where data was available.

#### Population data

In Bangkok, we initially placed a 1km x 1km grid cell over the central part of the city (337 grid cells) and estimated the population size using population size estimates from LandScan15. We also identified the grid cell for each of the Bangkok sequences. At the province-level we used population size estimates from the national census.

#### Call detail records (CDR)

To estimate adult mobility across provinces and within Bangkok, we used the call detail records (CDR) of over 11 million mobile phone subscribers between August 1, 2017 and October 19, 2017 from the third largest mobile phone operator in Thailand (N=11.4 million subscribers; 26% market share). These data are described in more detail elsewhere16. Briefly, each subscriber was assigned a daily “home” location based on their most frequently used cellular tower. Travel between locations was estimated by tabulating the subscriber’s home location on one day relative to the day before. The location-to-location transition probability matrix was estimated by using the average travel from location i to location j (weighted by the population at location i), and normalizing travel such that the sum of travel from location i is equal to 1.

#### Aedes aegypti abundance estimates

We used previously published estimates of the probability of Aedes aegypti presence for 5km x 5km grid cells around the globe17. These estimates were generated by incorporating information on temperature, rainfall, vegetation indices from satellite imagery and fitting models to a large dataset of Aedes occurrence records. The fitted models were then used to predict elsewhere. For Bangkok, we extracted the *Aedes aegypti* estimate using the centroid of each grid cell. For the province level, for each province, we used the average across all the raster cells from the Aedes map that were contained within that province.

### Generation time distribution for dengue

To estimate the generation time distribution for dengue, we combined data on the incubation period, extrinsic incubation period and the lifespan of the *Aedes aegypti* mosquito as has previously been used for chikungunya^18^.

#### Human incubation period (HI)

We used a truncated log-normal distribution with a mean of 5.6 days and a standard deviation of 1.41 days and a maximum time of two weeks ^19^.

#### Human to mosquito transmission (HM)

Based on estimated durations of viremia, we used a truncated exponential distribution with a mean of 4.5 days and a maximum period of seven days ^20^.

#### Mosquito infectiousness (MI)

The period of mosquito infectiousness depends on the mosquito lifespan and the extrinsic incubation period. The average daily probability of survival for *Aedes aegypti*, has been estimated at 0.87 for up to 30 days ^21^, equivalent to a mean lifespan of 7.2 days. The extrinsic incubation period has been estimated at 6.1 days ^22^. To calculate the period of mosquito infectiousness, we initially draw the mosquito lifespan (MLS) using a truncated exponential distribution with a parameter of 7.2 days and a maximum value of 30 days. Next, we draw the age at which the mosquito gets infected (MAI) from a uniform distribution between 0 and the lifespan of the mosquito. Next, we draw the extrinsic incubation period (EIP) as a random exponential distribution with mean of 6.1 days. The total period of mosquito infectiousness (MI) is then equal to MLS - MAI – EIP. Values of MI less than 0 were considered unsuccessful onward infections.

#### Generation time distribution

We derived the empirical distribution of the generation time by simulating 10,000 values for HI, HM and MI and summing them. Individuals who are viremic for longer are more likely to infect mosquitoes. Similarly, mosquitoes that are infectious for longer are also more likely to infect more individuals. We therefore weighted the probability of each generation time by the length of HM multiplied by the length of MI. We obtained a mean generation time of 18.2 days and a variance of 37.2 days, which we approximated using a gamma distribution with the same mean and variance (Figure S1).

### Time resolved phylogenetic trees

Taking each serotype in turn, we aligned the full genome sequences using the Muscle algorithm in MEGA2^23^. We built Bayesian time-resolved phylogenetic trees using BEAST 2.5.0^11^. We used a strict clock, a General Time Reversible nucleotide substitution model, as determined by jModelTest2^24^, and a Bayesian skyline prior. Similar coalescence times were found using a relaxed clock.

### Probabilistic model

#### Overall inferential strategy

We use a likelihood-based approach to model the probability of the observed location of pairs of sequences in a time-resolved phylogeny. We initially use the time-resolved phylogenies and information on the generation time distribution to estimate the number of generations that separates each member of a pair of sequences in the phylogeny from their Most Recent Common Ancestor (MRCA). This then allows us to consider single transmission generations rather than overall viral flow. We develop models of viral movement between each location in our study area (whether it was sampled or not) for each transmission step. These viral movement models incorporate estimates of human mobility with the potential for differences in movement for cases compared to the susceptible population, as well incorporating effects of time-varying serotype-specific immunity and vector distributions. We allow for infection to occur at the infector’s home location (which requires the infectee travelling to the infector’s home location), the infectee’s home location (which requires the infector to travel to the infectee’s home location) or in a tertiary location (where both parties would have to travel there).

We use the viral movement matrix for single transmission to calculate the viral movement after *G* generations via matrix multiplication. This approach integrates over all possible pathways that links two locations. To specifically incorporate observation processes, we consider the probability of sequencing an infection at each space-time unit.

To inform this model, we use an integrative approach that brings in detailed data from mobile phone operators that captures how people move and interact with each other, maps that estimate how populations are distributed, maps on vector suitability, and the long-term spatio-temporal distribution of serotypes.

#### Notation

For a pair of cases, *C*_*A*_ and *C*_*B*_, in a phylogenetic tree: *C*_*A*_ has home location *L*_*A*_, was sick at time *T*_*A*_ and has sequence *Seq*_*A*_; case *C*_*B*_ has home location *L*_*B*_, was sick at time *T*_*B*_, and has sequence *Seq*_*B*_. The time of the MRCA between *C*_*A*_ and *C*_*B*_ is *T*_*m*_ and the location of the MRCA is *L*_*m*_. We denote *G*_*A*_ and *G*_B_ the number of transmission generations that separate *C*_*A*_ and *C*_*B*_ from their MRCA. *Obs*_*LiTA*_ is 1 if a sequence was observed at location *Li* at time *T*_*A*_ and 0 otherwise. *Obs*_*LiTB*_ is defined in a similar manner for time *T*_*A*_.

### The single transmission generation matrix

Initially let us consider a single transmission generation. The probability that two individuals, *i* and *j*, are in the same location and are in contact (via a mosquito) given *i* lives in location *a* and *j* lives in location *b* can be written down as:

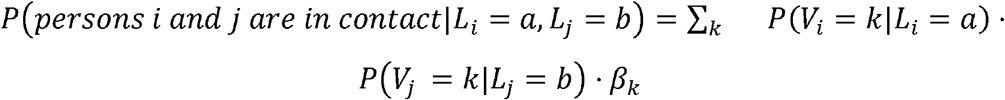

Where *P*(*V*_*i*_ = *k*|*L*_*i*_ = *a*) is the probability of individual *i*, whose home location is in *a*, visiting location *k* and *P*(*V*_*j*_ = *k*|*L*_*j*_ = *b*) is the probability that individual *j* also visits location *k* and *β*_*k*_ is the location specific probability of transmission.

At time *τ*, one infector *i* that lives in location *a* is expected to transmit to the following number of persons living in location *b*:

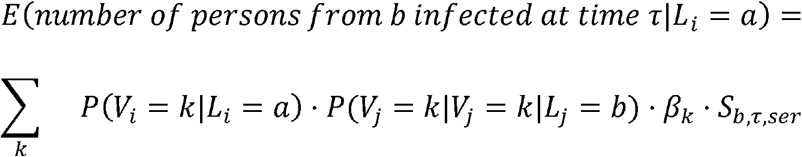

where *s*_*b,τ,ser*_ is the number of susceptible people to serotype ser living in location *b* at time *τ*. The total expected number of persons infected by infector is *i*:

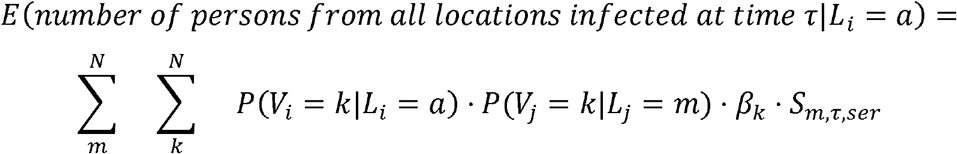

Conditional on transmission occurring, the probability that the infectee has home location in cell *b* is the ratio of these terms:

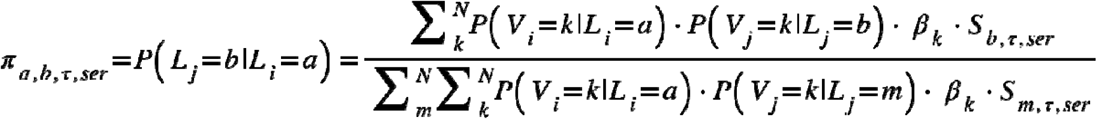

We can create a N x N transmission matrix,, where *N* is the total number of locations, that sets out the transmission probabilities between all pairs of locations at a point in time for a single transmission. The element *[a,b]* of the matrix is.

#### Characterizing human mobility

We use mobile phone data to characterize human mobility. Initially, we extract a matrix from the CDR data that sets out the probability that an individual that lives in location *a* visits location *k*.

CDRs come from adults, whereas dengue is concentrated in children, who are potentially more likely to spend more of their time at home. In addition, sick individuals may travel differently than healthy individuals and spend more time at home. In this way, the mobility of infectors may differ from the susceptible information.

To allow for different periods at home for susceptible individuals compared to those in the CDR data and to assess whether there exists differential mobility by illness status, we incorporate separate parameters for the probability of being at home for infectors and the susceptible population (,) to reflect additional time at home compared to that extracted from the CDR data.

For the home cells of infected individuals:

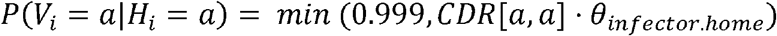

For the home cells of the rest of the population:

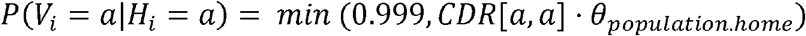

In each case, for non-home cells, we rescale the probabilities so that the sum of all movements remains equal to 1.

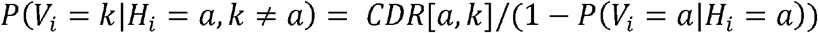

Where *CDR* [*a, k*] reflects the movement probabilities from the CDR data.

#### Factors affecting transmission

As there may be factors that allow for different probabilities of transmission across locations, we allowed for differential probability of infection by location based on the mosquito presence in that location.

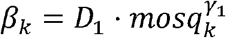

Where parameter *γ*_l_ is to be estimated, *mosq* represents the estimated suitability for *Aedes aegypti* mosquitoes in location *k. D*_l_is a proportionality constant (which gets cancelled out).

#### Factors affecting susceptibility

The number of susceptible individuals living in a location will depend on the level of historic infection in that location, in a potentially time-varying serotype-specific manner.

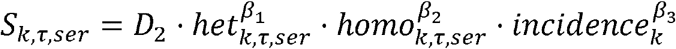

Where *het*_*k, τ,ser*_ is the incidence of cases caused by different serotypes in the two prior years in location *k*; *homo*_*k, τ,ser*_ is the incidence of cases caused by the same serotype in the two prior years in location *k; incidence*_k_ is the incidence of all cases over the study period in location k. For the Bangkok analyses, we use the serotype specific geolocated case data. For the nationwide analysis we use the national reporting system from the Ministry of Public Health (MOPH). The national MOPH system is not serotype specific, however, a small number of cases from all around the country are serotyped each year by the national reference center in Bangkok. To obtain serotype-specific incidence estimates for each year, we multiplied the proportion of cases that came from each serotype within each province-year by the overall number of cases for each province-year. Where there were no serotyped cases for a province-year, we used the closest province where there were serotyped cases.

#### Probability of virus being within each location after G transmission generations

To calculate the probability of the home location being within location *k* after *G* transmission generations, we can use matrix multiplication that integrates over all possible pathways connecting two locations.

Where *t*_*l*_ is the time of generation *G*_*l*_.

#### Probability of observing a pair of cases in two specific locations

Conditional on sequences being observed in location *L*_*A*_ at time *T*_*A*_ and *L*_*B*_ at time *T*_*B*_, the probability that *C*_*A*_ has home location *L*_*A*_ and *C*_*B*_ has home location *L*_*B*_ can be written down as:

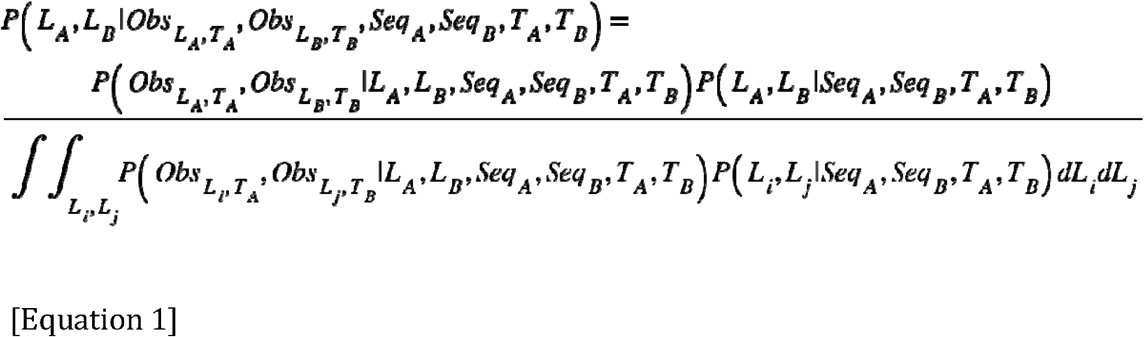

We can consider that the location of the two cases is dependent on the location of their MRCA and the number of transmission generations that separate them from the MRCA.

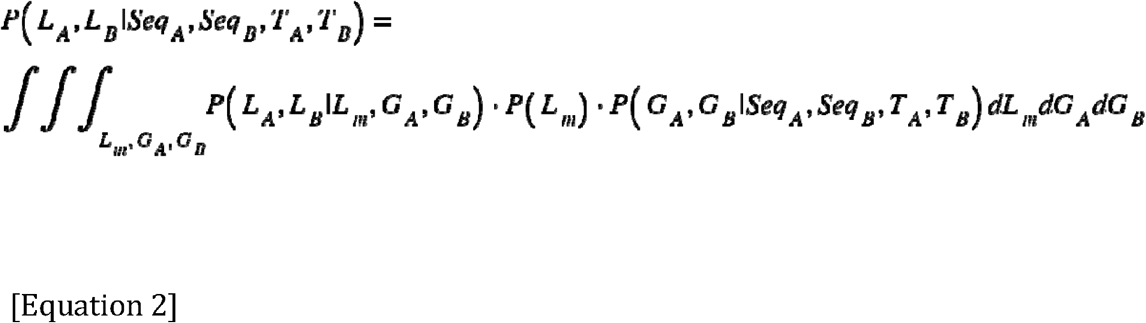

We consider that the observation processes across locations are independent of each other. In addition, each transmission event is considered independent of other transmission events. The probability of observing a case at location *L*_*i*_ at time *T*_*A*_ does not depend on the location of the MRCA or the number of generations separating the case to the MRCA. We can also substitute in Equation X2 into Eq. X1. Finally, we consider discretized space – either 337 1km x 1km grid cells throughout central Bangkok or the 76 provinces of Thailand.

Equation 1 therefore becomes:

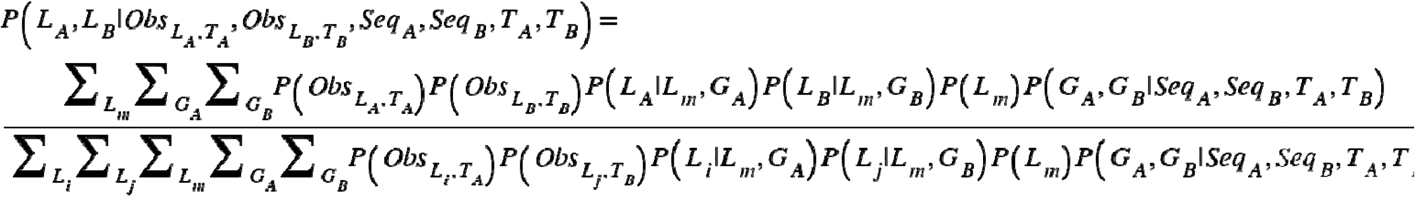

#### Probability of G generations between the MRCA and a case

We can extract the joint probability that case *C*_*A*_ is separated from the MRCA by *G*_*A*_ transmission generations and case *C*_*B*_ is separated from the same MRCA by *G*_*B*_ transmission generations using the generation time distribution, for dengue and the time-resolved phylogenetic tree.

If we assume that the generation time distribution is gamma distributed with parameters and and that all transmission events are independent of each other, the sum of *g* gamma distribution is also gamma distributed with parameters *g* and. In addition, from a genealogy,, we can extract the evolutionary time, *E*_*A*_, separating *C*_*A*_ from the MRCA and *E*_*B*_, separating *C*_*B*_ from the MRCA. We can therefore estimate the probability of *g* transmission events as follows over many trees as follows:

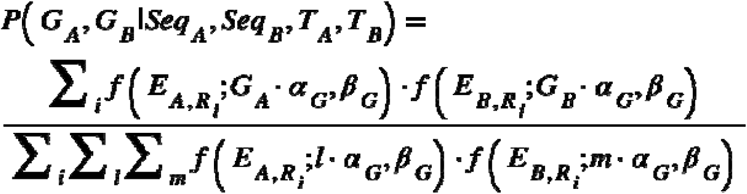

As any spatial signal will be heavily diluted after many transmission generations, to optimise computational performance we restrict our analyses to pairs where the mean estimated number of transmission generation is <25, we perform a sensitivity analysis where this is extended to 40 generations with very similar results (Figure S3).

#### Observation probability ()

We cannot know the true number of infections occurring within each space-time unit. Given the long-term endemicity of dengue in the region, we assume that the number of infections will be approximately proportional to the size of the population within each location. Therefore, the probability of observation (the probability of sequencing the virus causing an infection event) at location *k* at time point *t* is approximately proportional to the number of sequenced viruses from that year and location for that serotype divided by the size of the population in that location.

We conducted a sub-analysis where we assumed unbiased observation. In this analysis we assumed that the probability of observation was 1 across all space-time locations, we obtained very different results (Figure S6).

We further assessed the performance of this approach using a simulation model that imposed a heavily biased observation process. Our inference framework was able to correctly identify all parameters (Figure S7, see below for simulation model details).

#### The location of the MRCA (P(L_m_))

The probability of the MRCA for each pair of cases will depend on the long-term history of dengue in the communities, which cannot be estimated using the presented approach. Instead we assume that *P*(*L*_*m*_) is proportional to the size of the population in that location. To assess the sensitivity of this assumption, we conducted a separate analysis where *P*(*L*_*m*_) was assumed to be the same across all locations, with identical results (Figure S4). This suggests that we do not need to probabilistically assess where the start point is for the MRCA that links two cases in a phylogeny.

#### Likelihood

We can calculate the likelihood using all pairs of available sequenced viruses as follows:

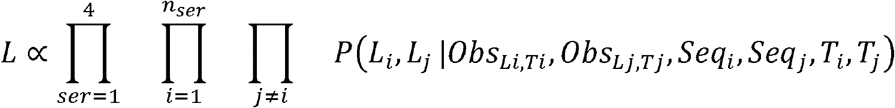

Where *n*_*ser*_ are the number of sequences available from serotype ser.

#### Identifying the Maximum Likelihood Estimate

We use a maximum likelihood approach to estimate the parameters linked to the mobility, transmission and susceptibility (*θ*_*home*.*sick*_, *θ*_*home*.*population*_, *β*_*1*,_ *β*_*2*_, *β*_*3*,_ *γ*_*1*_) We identify the Maximum Likelihood Estimate using an unconstrained non-linear quasi-Newton optimization approach25.

In order to incorporate uncertainty, we use a bootstrapping approach where we randomly sample all the available sequences with replacement over 100 iterations and recalculate the maximum likelihood estimate for each parameter each time. The 95% confidence intervals are then taken from the 2.5 and 97.5 percentiles of the resulting distribution.

#### Using fitted values to estimate patterns of viral flow at each transmission generation

Once we have fitted values for the parameters, we can calculate the *П* _*τ ser,gen*_ matrix for each month between 1994 and 2012, each serotype, and each transmission generation. From these matrices, we can extract the probability that the virus is in each location given a specified serotype, location and time of introduction and number of generations. From these matrices, we calculate the cumulative distribution function of the distance between where the virus started and where it is after different numbers of generations, averaging over time and serotype. We compare this to the cumulative distribution function of how far cases are from their home at any time. This highlights that viral mobility requires both movement of cases and the susceptible population.

We also calculate the mean proportion of times that the most likely (non-home) destination is the same across serotypes. This allows us to assess whether viruses across the serotypes take the same routes or whether serotype-specific immunity changes the most likely pathways.

#### Model fit

In order to assess model fit, we perform held out validation. In Bangkok, we remove all sequences from 10% randomly chosen locations, we then refit the model and then estimate the probability of observing sequences in the locations not included in the model fitting process. For the nationwide analysis, as we have fewer locations with sequences available, we undertake the same process but hold out a single province in turn.

For incremental windows of probability between 0 and 1, we identify all location-years where a virus was predicted to have been observed within the held-out locations. We then calculate the mean proportion of times a sequence was observed within those identified location-years.

#### Estimates of spread using a transmission simulation

Using the fitted parameter values we conduct a forward simulation at both the Bangkok and province levels. Taking each month between 1994 and 2012 and each serotype in turn we apply the following algorithm:

i. Randomly introduce a single infection in one location where all locations have the same probability of being the source.
ii. We generate daughter infections from the index using a random draw from a Poisson distribution with mean *Reff* (representing the effective reproductive number).
iii. We identify the location for each daughter infection using a random draw where the probability of each location is taken from the *П* _*τ ser,gen*_ matrix.
iv. Repeat (ii) and (iii) for 20 generations.
v. Repeat (i)-(iv) 50 times

For each iteration, we calculate the average number of locations that have had at least one infection at each generation, average over all time points and all serotypes.

To assess the impact of mobility patterns and immunity on the number of locations affected, we repeated the analysis with the following adjustments:

> Scenario a: All initial introductions were in the most connected location only (as defined as the location with the lowest probability of staying within your home location).
>
> Scenario b: All initial introductions were in the least connected location only (as defined as the location with the highest probability of staying within your home location).
>
> Scenario c: No difference in the mobility of the cases as compared to the susceptible population. This was achieved by forcing the *θ*_*infector*.*home*_ parameter to be the same as the fitted *θ*_*population*.*home*_ parameter.
>
> Scenario d: Susceptible population mobility is equal to that of the adult population. This was achieved by forcing the *β*_*population*.*home*_ and *θ*_*infector*.*home*_ parameters to be zero.
>
> Scenario e: No impact of immunity. This was achieved by forcing the *β*_l_ and *β*_2_ parameters to be zero.
>
> For each scenario, we calculated the proportion of locations affected at each generation and the relative number of locations affected compared to the base model. We also conducted sensitivity analyses where the R_eff_ was varied from 1.3 to 1.1 and 1.6.

### Age mixing model

We use an equivalent approach to characterize the age-dependent mixing of the population. Instead of considering the probability of the virus transitioning between two locations, we consider the probability of transitioning between individuals of two ages.

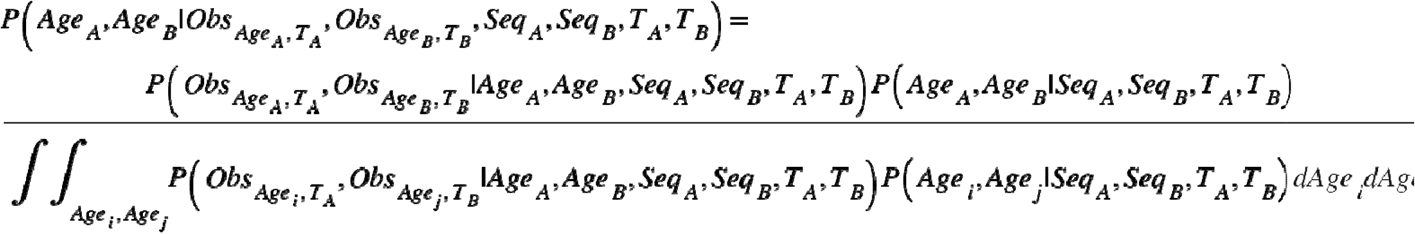

We can consider that the age of the two cases is dependent on the age of the MRCA and the number of transmission generations that separate them from the MRCA.

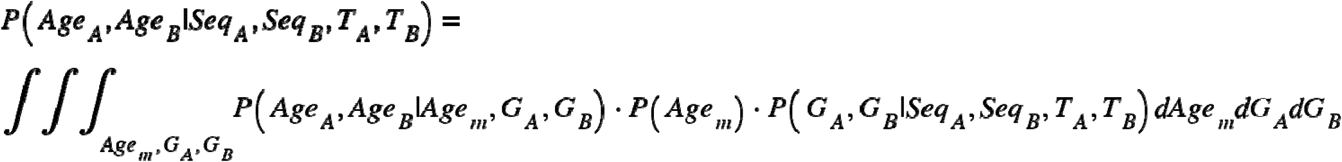

We consider that the observation processes across ages are independent of each other. In addition, each transmission event is considered independent of other transmission events. The probability of observing a case at age *Age*_*i*_ at time *T*_*A*_ does not depend on the location of the MRCA or the number of generations separating the case to the MRCA. We can also substitute in Equation X2 into Eq. X1. Finally, we consider discretized ages.

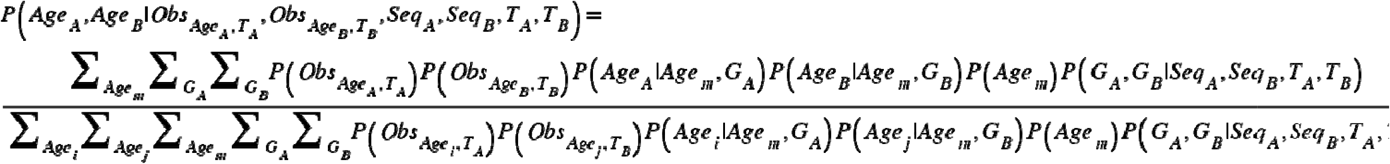

#### The age transition matrix

Initially let us again consider a single transmission generation. We assume that the age-specific susceptibility of the population is stable over time and that the probability of exposure does not differ by age group or serotype. The probability that two individuals, *i* and *j*, of ages *Agei* and *Agej* are in contact (via a mosquito) can be written down as:

Where is the age specific probability of contact between individuals of ages *a* and *b*.

The expected number of infected persons coming from individuals of age *b* conditional on an infector, *i*, being of age *a* is:

Where is the number of susceptible people of age *b*.

The expected number of infected persons coming from individuals of all ages, conditional on an infector, *i*, being of age *a* is:

Conditional on one transmission generation where the infector *i* is of age *a*, the probability of the infectee with age *b* is therefore the ratio of these terms. We define this probability as *ϕ*_*a,b*_.

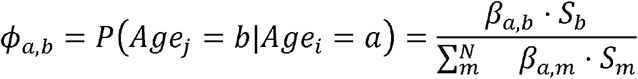

We can create an *N*_*age*_ x *N*_*age*_ transmission matrix, Ф, that sets out the transmission probabilities between all ages for a single transmission. Where element *[a,b]* of the matrix is *ϕ*_*a,b*_. We use a maximum age of 70 years.

#### Age contact matrix

To parametrically characterize the age contact matrix, we use an discretized exponential decay parameter, *θ*_*age*_, that captures the probability that two people interact, as a function of the difference in their ages, such that *β*_*a,b*_ = *f* (*a* -*b*; *θ*).

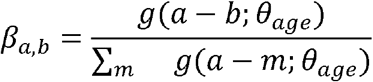

Where *g*(*x*; *θ*_*age*_) = *θ*_*age*_ · *exp* (-*θ*_*age*_ · *x*).

#### Age specific susceptibility

To characterize the susceptibility, we assume that the probability of being susceptible is equal to 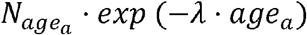,where 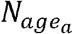 is the number of people of age *a* in the national census and the force of infection, *λ*, is assumed to be 0.04 26. We conduct a sensitivity analysis where the force of infection is varied to 0.02 and 0.06 with unchanged results (Figure S5).

#### Observation probability (P(Obs_Agei,TA_))

We assume that the probability of observing a case of age a is proportional to the number of sequenced viruses of that age for that serotype divided by the estimated size of the susceptible population of that age (Sa).

#### Likelihood for the age model

We can calculate the likelihood using all pairs of available sequenced viruses as follows:

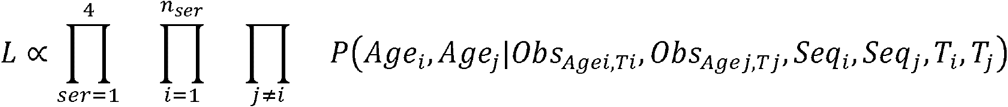

Where *n*_*ser*_ are the number of sequences available from serotype ser.

We use a maximum likelihood approach to estimate the parameter *θ*_*age*_. In order to incorporate uncertainty, we use a bootstrapping approach where we randomly sample all the available sequences with replacement over 100 iterations and recalculate the maximum likelihood estimate for each parameter each time. The 95% confidence intervals are then taken from the 2.5 and 97.5 percentiles of the resultant distribution.

Once we have fitted the *θ*_*age*_ parameter, we calculate from the matrix of *ϕ*_*a,b*_, the proportion of transmissions that are between individuals that have <2y in age between them. We use an equal weight for the age of the infector across all ages and only consider individuals between the ages of 1 and 15 as they represent the majority of the susceptible population. We compare this to the scenario where all individuals have the same probability of contact, irrespective of age (i.e., *β*_*a,b*_ 1/70, for all *a* and all *b*), which is the minimum possible value. We also calculate the equivalent value using the POLYMOD contact matrix, an empirical estimate of age-mixing patterns in European countries.

### Simulation study

In order to ensure that our model is able to correctly identify parameters, we built a simulation framework with known parameters using 50 randomly selected grid cells from the Bangkok dataset. Where the population size, the historic incidence and the probability of between cell movement were taken from the observed data. As the observed heterogeneity in mosquito presence was limited in Bangkok (Figure S2), we simulated mosquito presence in each location using a Uniform distribution between 0 and 1. For the recent heterotypic and homotypic cases we used a randomly selected time point from the observed distributions of cases and assume all cases came from serotype DENV1.

We fixed the parameter values as follows:

– Additional time being at home for susceptible population compared to adults (*θ*_*susceptible*.*home*_) (logit scale): -0.5
– Additional time being at home for cases compared to adults (*θ*_*infector*.*home*_) (logit scale): -0.05
– Mosquito exponent: 1.0
– All incidence exponent: 0.5
– Recent heterotypic incidence exponent: 0.3
– Recent homotypic incidence exponent: -0.3

We then calculated the *П* _*τser,gen*_ transmission matrix using these known parameters and simulated transmission events using the following algorithm:

1. Randomly select a starting location (H_0_) by randomly choosing a location, weighted by the population in that location. This will represent the MRCA case (C_0_) between the two observed viruses.
2. Draw the number of generations (*g*) between the MRCA and one of the observed isolates, where the number of generations is between 15 and 19 generations and the probability of 15 generations is 0.1, 16 generations is 0.2, 17 generations is 0.4, 18 generations is 0.2, 19 generations is 0.1.
3. For case C_0_ identify where they will transmit to (H_1_) using a random draw with the probabilities of each destination location coming from the H_0_ row of the *П* _*τser,gen*_ matrix.
4. Repeat step 3) g times using the destination of the previous step as the start location each time
5. Repeat steps 1-4 2000 times to generate 2000 pairs of cases
6. We assumed that the probability of observing (i.e. sequencing) the virus from a case was unequal across locations. The probability of observing a case at a location (*ρ*) is taken from a random uniform distribution (U(0,1)). We randomly select 500 pairs of cases where the probability of observation of each pair is the product of the probability of observation at each of the two locations.

Using the observed pairs, we then used our framework to estimate the parameters of the model. We repeated the simulation 50 times and report the mean and 2.5 and 97.5 percentiles of the distribution for each parameter estimate.

To assess the importance of incorporating sampling bias in our estimates, we repeated the inference on our simulated data but assumed that all space-time locations had the same, equal chance of being observed.

## Ethics statement

This study was approved by the ethical review boards of Queen Sirikit National Institute of Child Health, and Walter Reed Army Institute of Research and the University of Florida. Case data was obtained from the results of standard confirmatory testing for dengue and therefore did not require informed consent.

## Data and code availability

[R code used for the analyses will be made available using a publicly available GitHub link upon acceptance. GenBank references for the sequences are available in Table S1. Metadata for the sequences will also be made available on a publicly accessible GitHub link upon acceptance].

## Supplementary materials

**Table S1. GenBank accession numbers and dates and location for all sequences used in analysis**.

[Will be attached as csv on acceptance]

**Figure S1:**
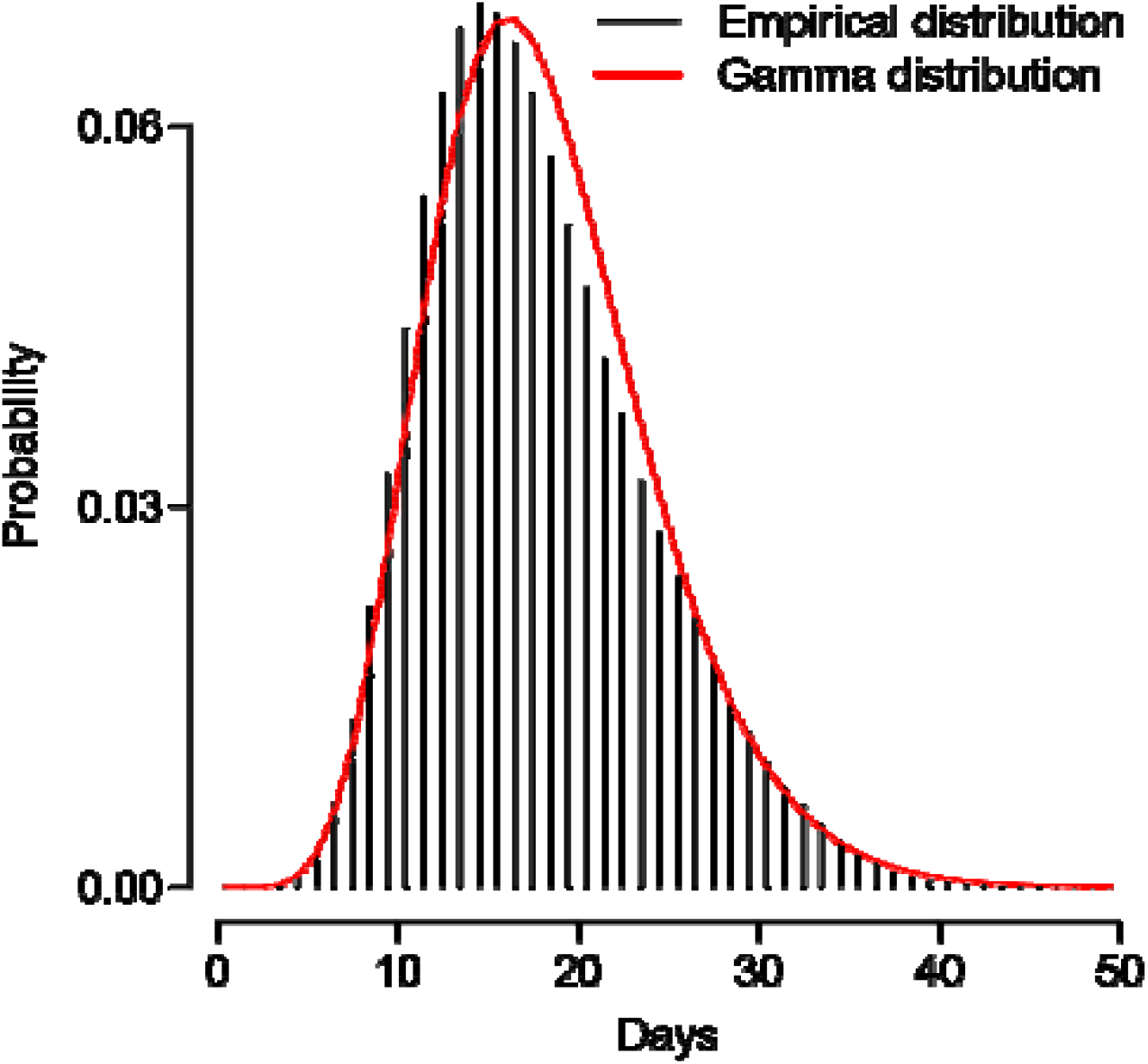
Generation time distribution for dengue. We compare the empirical distribution function (black) with that using a gamma distribution (red).

**Figure S2:**
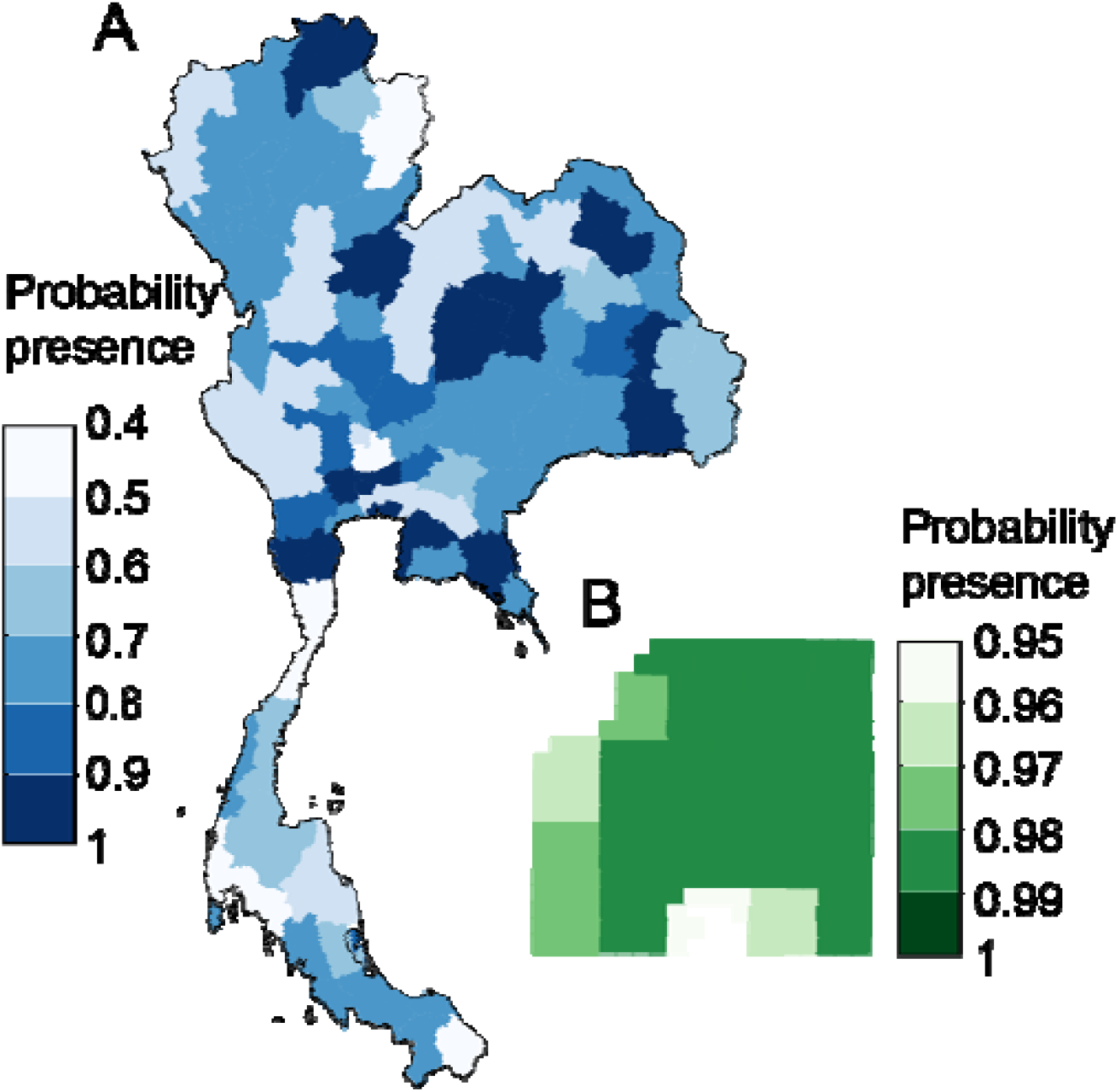
Aedes aegypti maps. **(A)** Mean estimated probability of Aedes presence across each province in Thailand. **(B)** Mean estimated probability of Aedes presence across each cell in central Bangkok. Note the different scales in the two locations.

**Figure S3:**
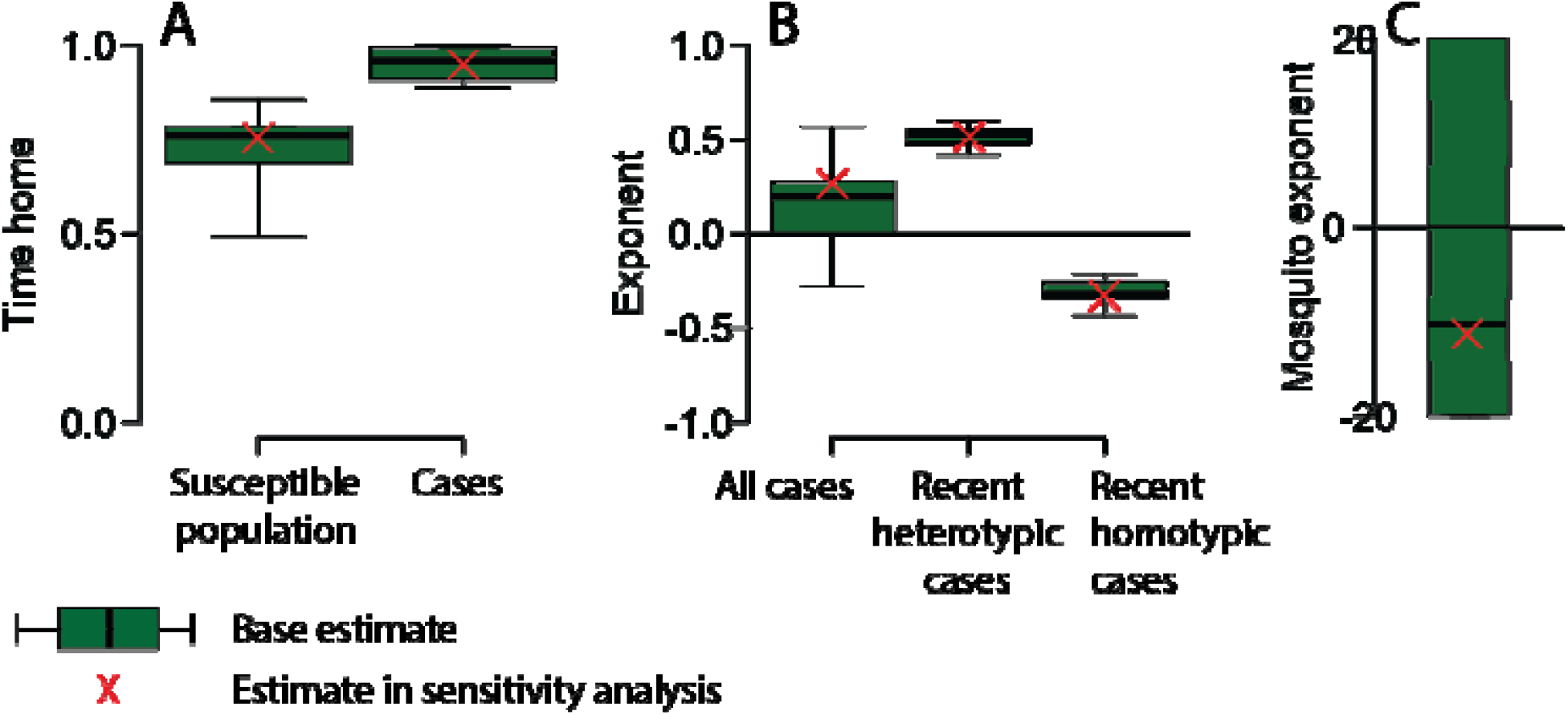
Sensitivity analysis where assume location of MRCA is equal across all locations. We compare the inferences made when we assume that the probability the MRCA is in any location is proportional to the size of the population within that location (base model, green) with when we assume all locations have the same probability (red cross). **(A)** Estimates for the time in home cell for the susceptible population and cases. **(B)** Exponents for all cases, recent homotypic cases and recent heterotypic cases and **(C)** exponent for the mosquito suitability estimate.

**Figure S4:**
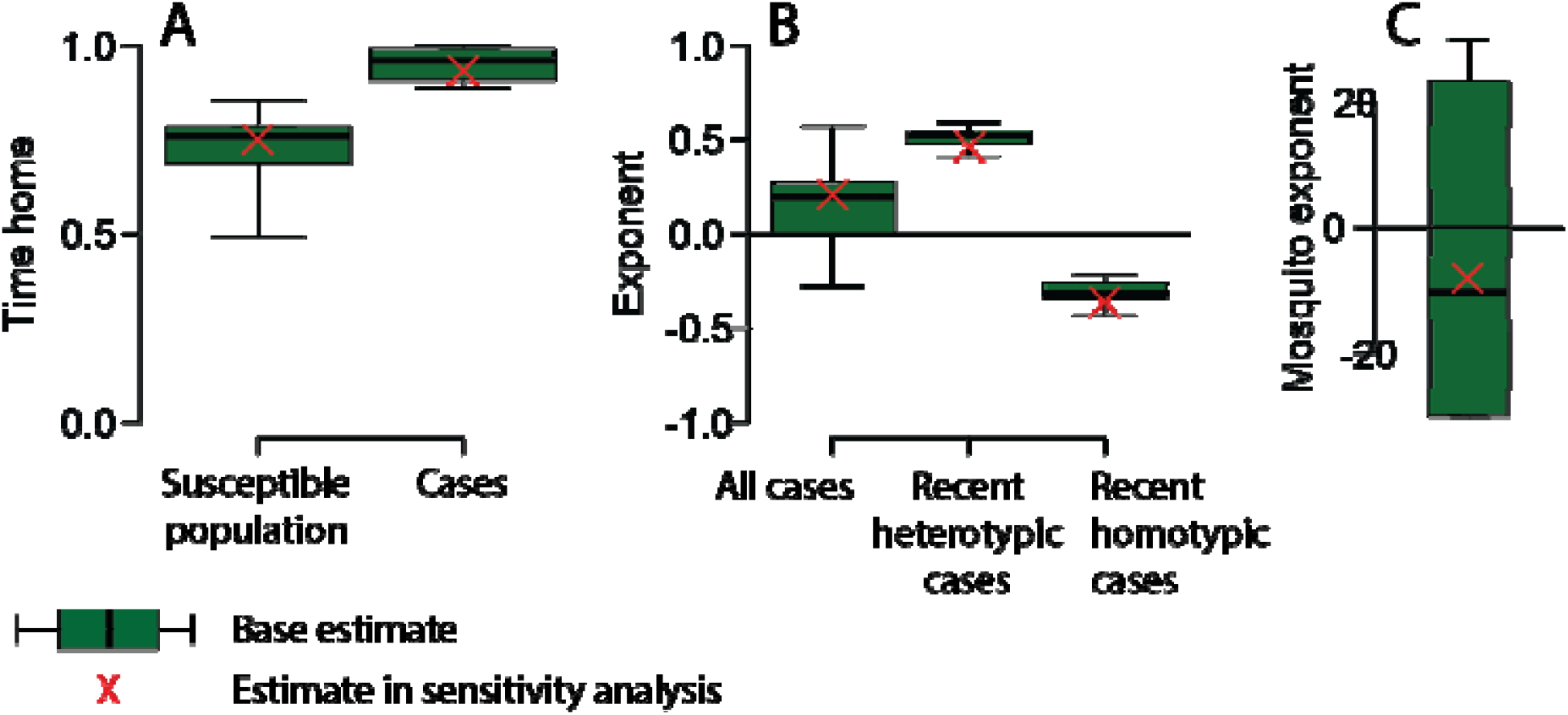
Sensitivity analysis where inference based on 40 generations. We compare the inferences made when we consider pairs of sequences where the expected number of transmission generations between the MRCA and each tip is <25 generations (base model, green) with inferences when we use a maximum number of 40 transmission generation (red cross). **(A)** Estimates for the time in home cell for the susceptible population and cases. **(B)** Exponents for all cases, recent homotypic cases and recent heterotypic cases and **(C)** exponent for the mosquito suitability estimate.

**Figure S5:**
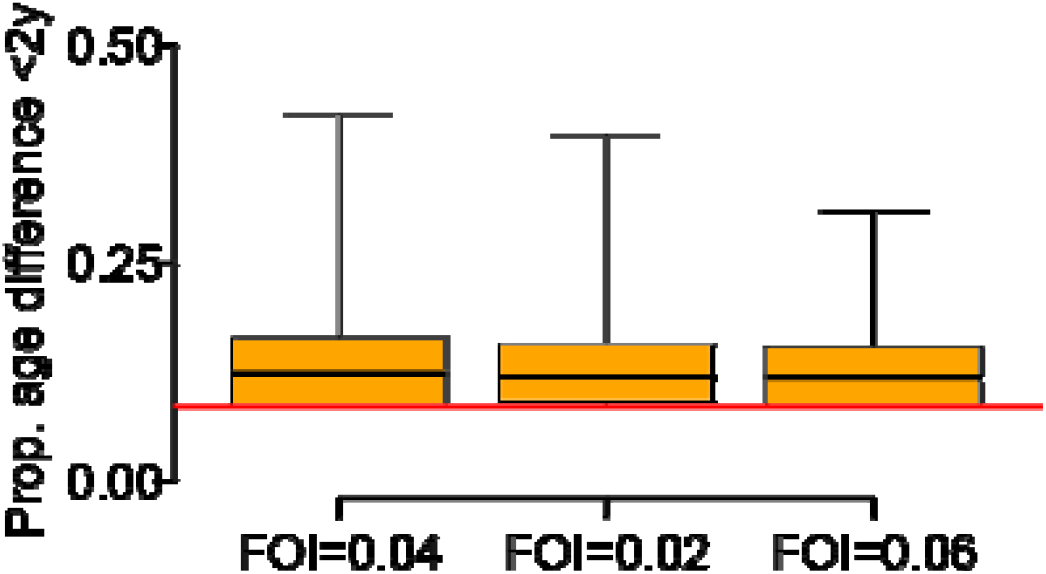
Sensitivity analysis where assume different force of infections. Estimates of the proportion of infections that are between individuals of within 2 years difference in age. The red line represents the scenario where there is no difference across all age groups and is the lowest possible value that can be taken.

**Figure S6:**
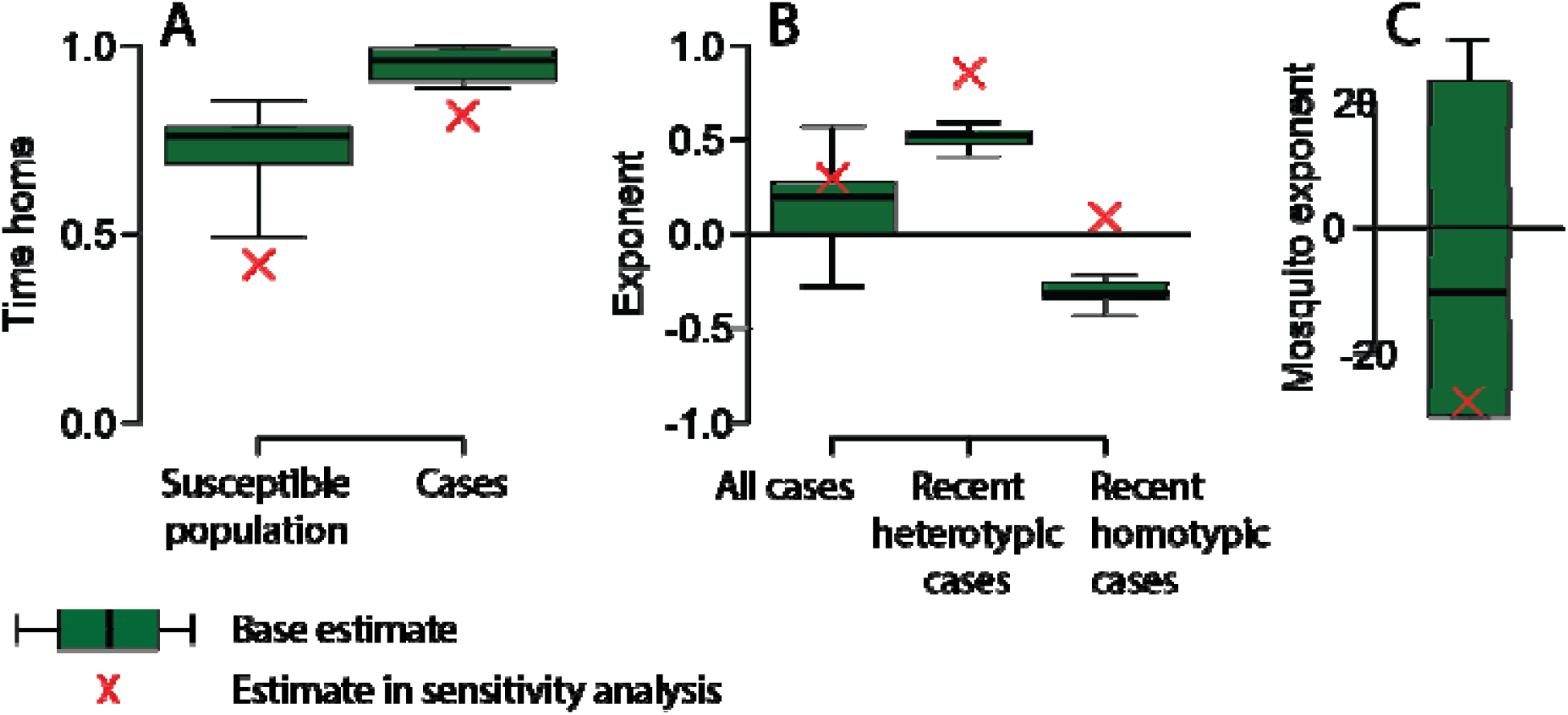
Sensitivity analysis where assume unbiased observation. We compare the inferences made when we account for sampling probability (base model, green) with when we assume even sampling in space and time (red cross). **(A)** Estimates for the time spent in the home cell for the susceptible population and cases. **(B)** Exponents for all cases, recent homotypic cases and recent heterotypic cases and **(C)** exponent for the mosquito suitability estimate.

**Figure S7:**
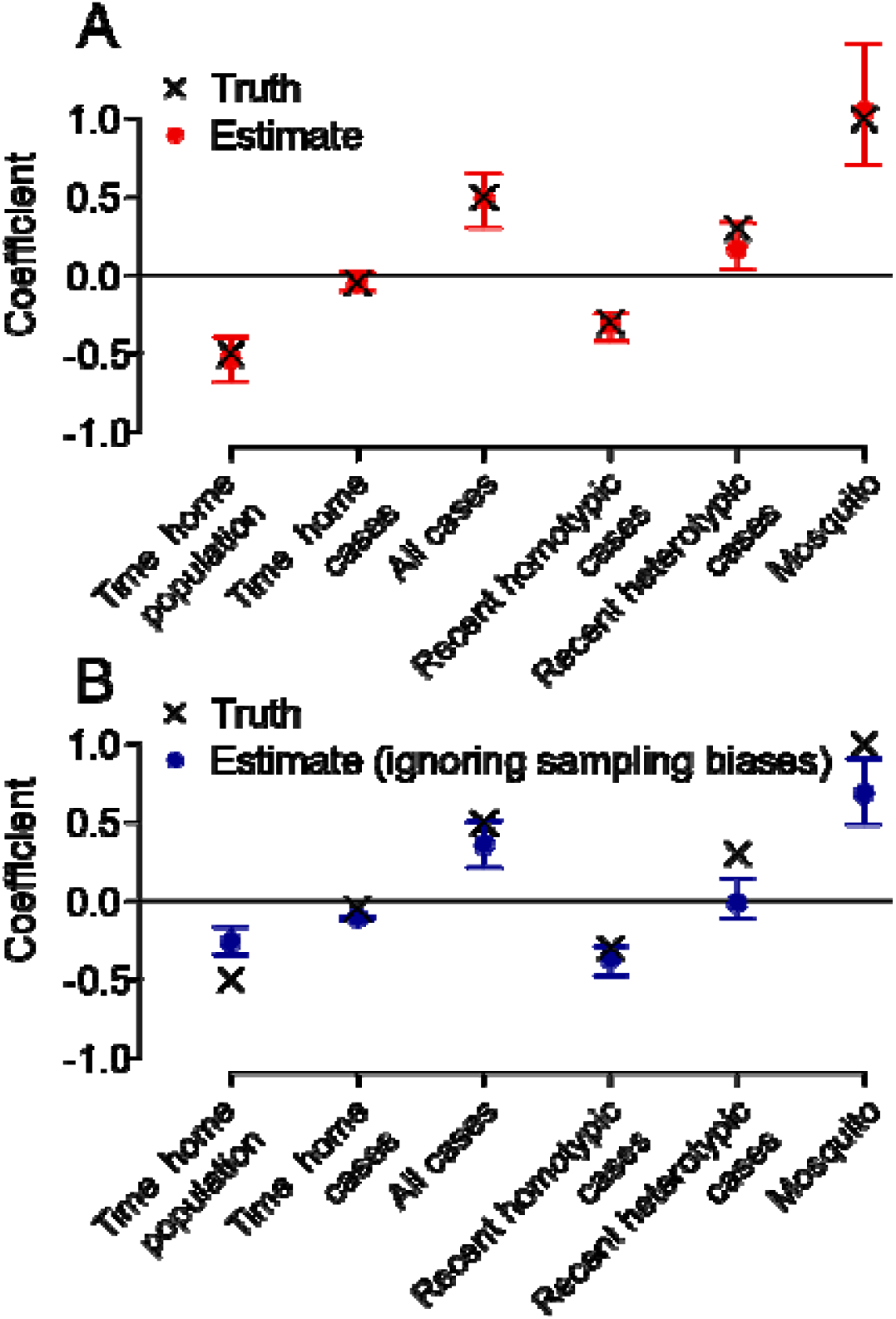
Performance of model using simulated data with known parameter values. **(A)** Comparison of mean and 95% range of estimates (red) from 20 simulations with true values (black cross) when appropriately accounting for spatiotemporal biases in reporting. **(B)** Comparison of mean and 95% range of estimates (blue) from 20 simulations with true values (black cross) when assuming equal probability of reporting in all locations.

**Figure S8.**
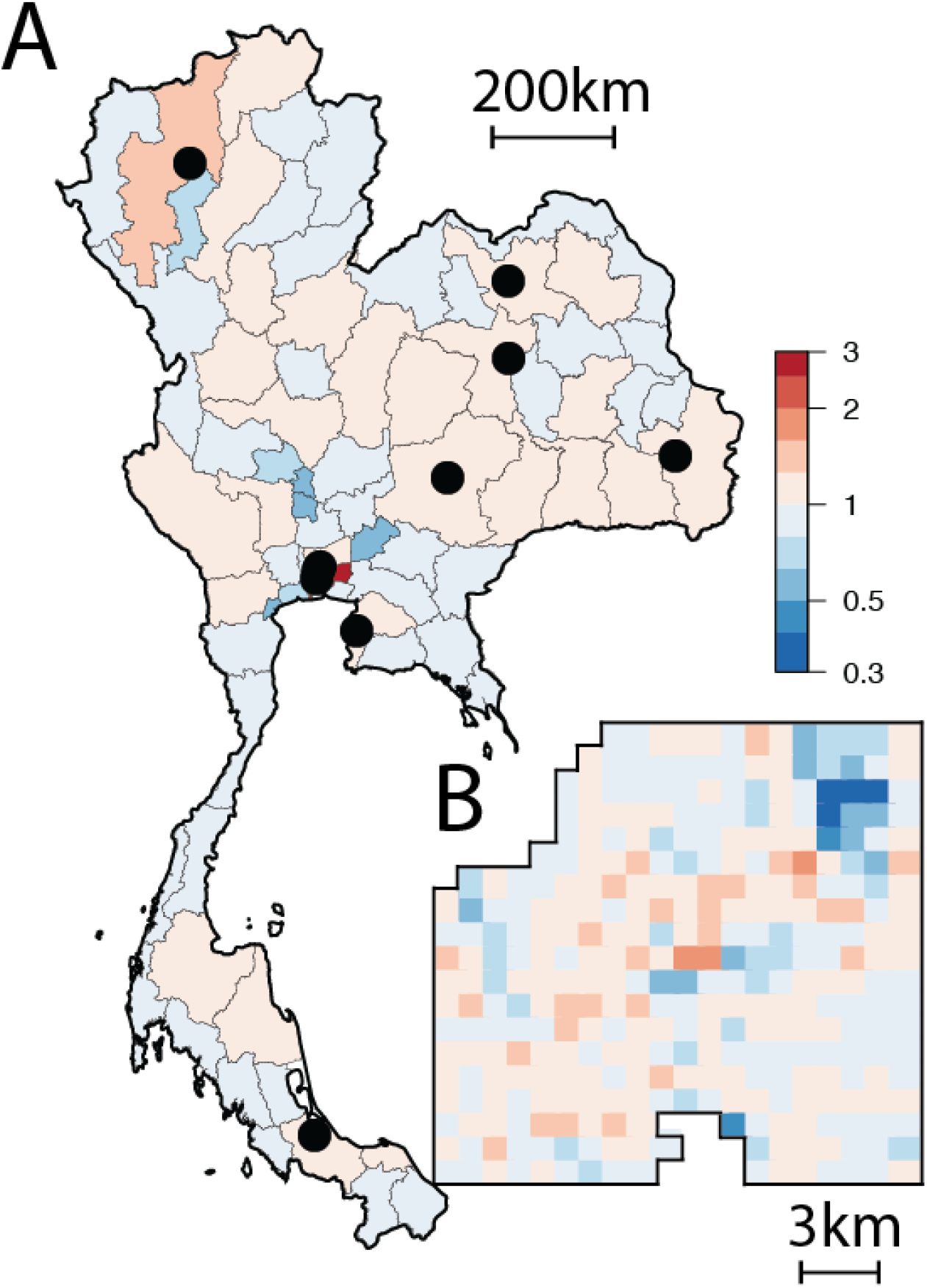
**(A)** Relative risk of movement of virus to each province compared to moving to a randomly selected province in a single transmission generation. The black dots represent the 10 largest cities in Thailand. **(B)** Relative risk of movement of virus to each grid cell within central Bangkok after a single generation.

**Figure S9.**
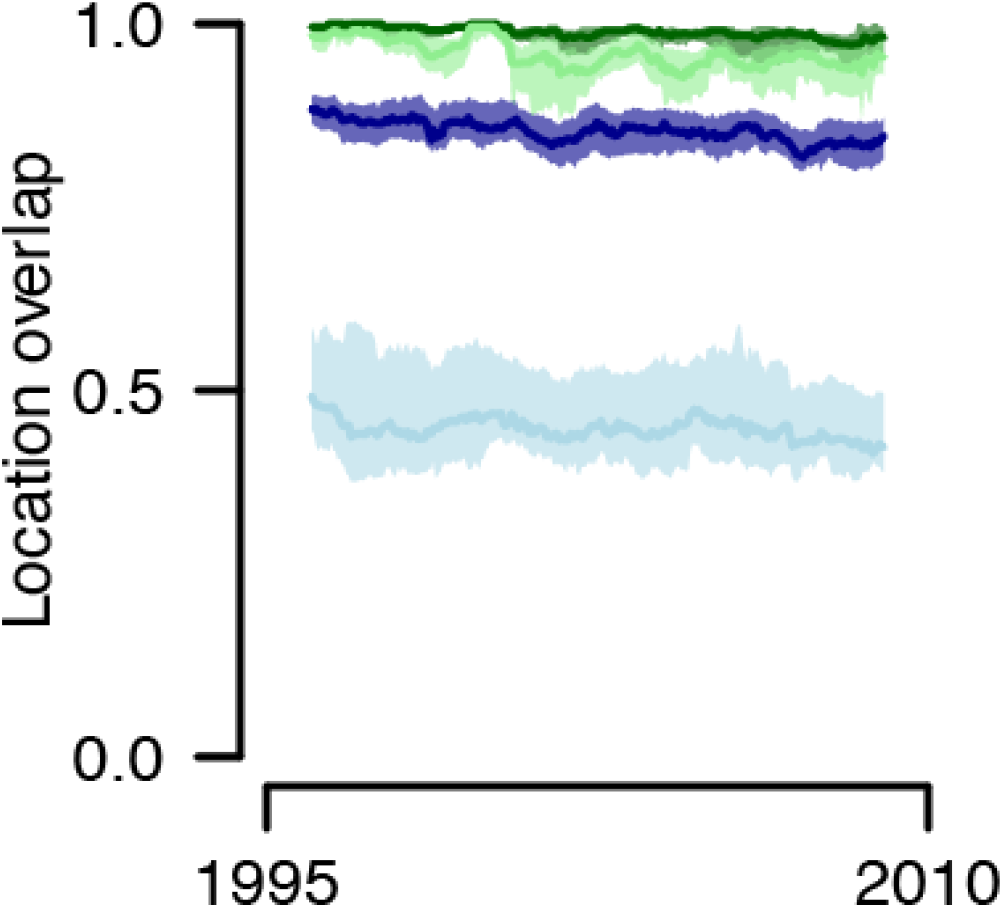
The probability of the likeliest non-home location being the same across serotypes in Bangkok and across provinces after a single transmission step (purple for Bangkok and dark green for provinces) and 20 generations (light blue for Bangkok and light green for provinces).

**Figure S10:**
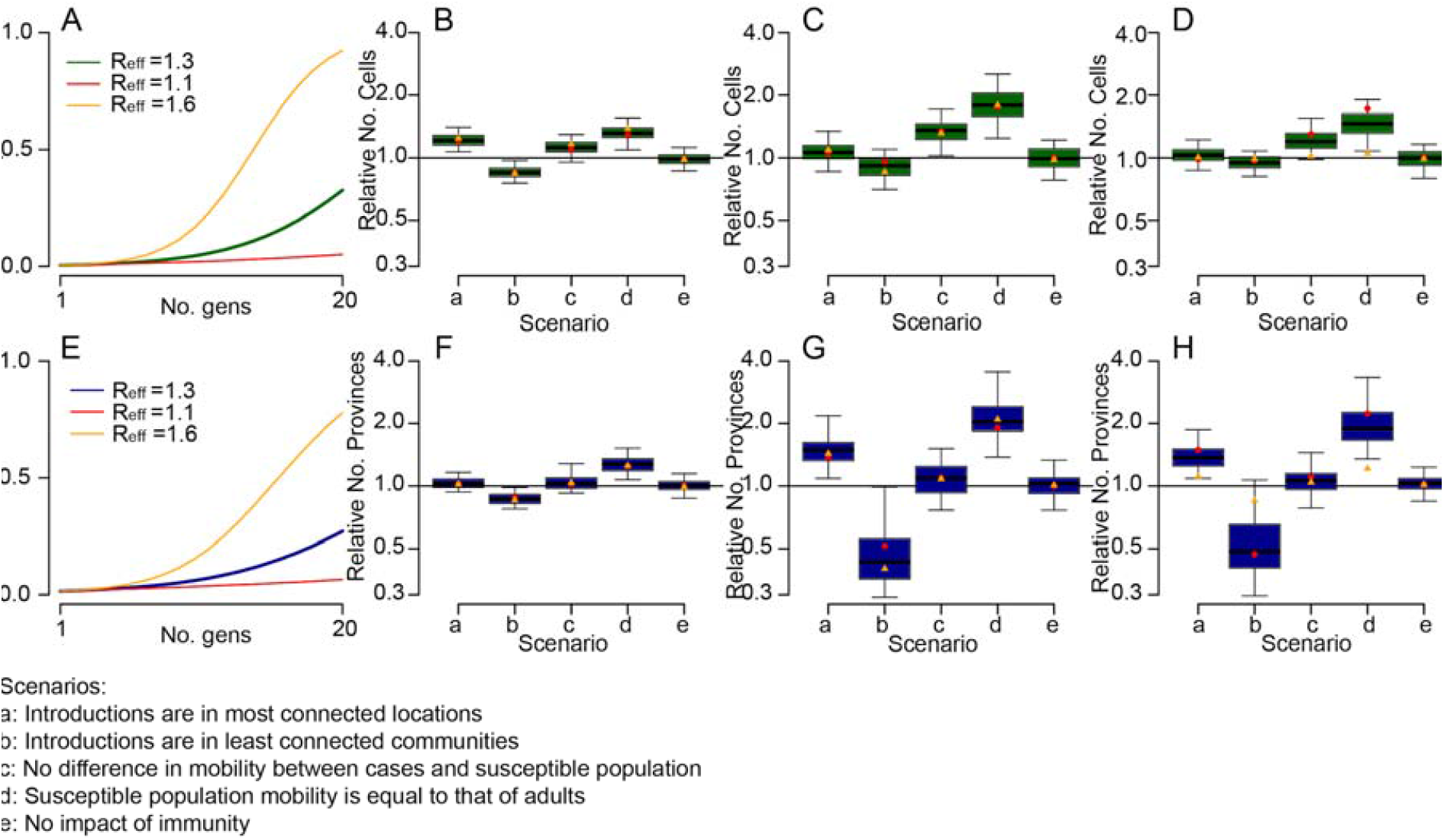
Sensitivity analysis where assume different R_eff_ on forward simulations. **(A)** The proportion of locations within Bangkok that have at least one infection, comparing different R_eff_. The number of Bangkok locations with at least one case for different scenarios relative to that in the base model after **(B)** one generation **(C)** 10 generations (approximately six months) and **(D)** 20 generations (approximately one year). **(E)** The proportion of Thai provinces that have at least one infection, comparing different R_eff_. The number of Thai provinces with at least one case for different scenarios relative to that in the base model after **(F)** one generation **(G)** 10 generations (approximately six months) and **(H)** 20 generations (approximately one year). The red and orange dots in panels **(B)-(D)** and **(F)-(H)** represent the scenarios with R_eff_ of 1.1 and 1.6 respectively and the green/blue boxes represent the scenario with an R_eff_ of 1.3 with an interquartile range and 95% confidence intervals.

